# Epidemiology of urinary tract infection among community-living seniors aged 50 plus: population estimates and risk factors

**DOI:** 10.1101/2024.08.16.24312133

**Authors:** Betsy Foxman, Marie Bangura, Neil Kamdar, Daniel M Morgan

## Abstract

**Background:** Urinary tract infection (UTI) is common in all ages but risk factors among adults 50 and older are not well studied. One unexplored potential risk factor is constipation, a known UTI risk factor among children.

**Objective:** Describe the UTI incidence among U.S. women and men aged 50 and older and the association between constipation and other risk factors and UTI.

**Methods:** A web interview was administered October 12-16, 2023 to U.S. adults aged 50 and older participating in a probability-based panel representative of the U.S. household population age 50 or older. The primary study outcome was self-reported healthcare provider diagnosed and treated UTI in the previous 12 months. All results were weighted to represent the U.S. household population.

**Results:** The 12-month UTI incidence was 19.8% among women and 6.4% among men. Recurrence was common: 10% of women and 7% of men with a UTI had 3 more UTIs in the previous 12 months. 32% of the population reported being constipated sometimes, frequently or always. UTI incidence increased with more frequent constipation in a dose response manner among women and men. After adjusting for age, gender, having a body mass index >30 and an overnight hospital stay in the previous 12 months, those reporting sometimes being constipated were 3.69 times, and those often or always constipated were 5.48 times more likely than those never constipated to have a UTI in the previous 12 months.

**Discussion:** This is the first report of an association between UTI and constipation among older adults. Reducing constipation might reduce UTI incidence among older women and men.

## Introduction

Urinary tract infection (UTI) is one of the most common bacterial infections. Every year 0.7% of physician office visits (Santo L & K, 2023) and 1.8% of emergency department visits (∼2.3 million visits) in the United States are for UTI (Cairns & Kang, 2022). Due to the effectiveness of antibiotic therapy, mortality from complicated UTI is low: in the United States the age-standardized mortality is estimated at 3.44 per 100,000 population (X. Yang et al., 2022). However, UTIs are increasingly resistant to antibiotics. In a study of 233,974 *Escherichia coli* isolates from 174,185 individuals with UTI collected between 2016 to 2021 at Kaiser Permanente Southern California almost half (48%) of all isolates were resistant to one or more drug classes; 11.9% were resistant to three or more drug classes (Ku et al., 2023).

The most recent population-based estimates of UTI prevalence and incidence in the United States used self-reported data from the National Health and Nutrition Examination Survey III (1988 to 1994) (Griebling, 2005b, 2005a). At that time, lifetime UTI prevalence was 53% and 14% for women and men, respectively. UTI incidence in the previous 12 months among women peaked at ages 18 to 24 at 27.1%, decreasing to 9.2% at ages 65 to 74, and rising to 11.8% at ages 85 and older. For men the incidence was 0.9% at ages 18-24 rising gradually with age and peaking at 7.7% at ages 85 and older. An analysis of over one million outpatients with UTI treated at Kaiser Permanente Southern California between 2008 to 2017 showed that half of the UTIs among men occurred among those aged 65 and older compared to 23% of UTIs among women. (Bruxvoort et al., 2020). In 2022, 16% of the U.S. population was 65 years of age or older [https://www.neilsberg.com/insights/united-states-population-by-age/]; UTI-related spending is estimated to represent about one-third of the total annual Medicare costs for patients diagnosed with UTI (Sulham & Hammelman, 2021).

Clinically, the symptoms of uncomplicated UTI among those over 65 include those found among younger individuals: frequency, urgency, and dysuria [Gharbi et al. 2019]. However, post-menopausal women are more likely than pre-menopausal women to report generalized symptoms, including abdominal pain and constipation and less likely to report urinary frequency and painful urination (Arinzon et al., 2012). Among women and men aged 65 and older with UTI, confusion, malaise, enuresis, and urinary incontinence also are common (Gharbi et al., 2019). In addition, older individuals are more likely to acquire UTI when in the hospital – even when no devices are present (Strassle et al., 2019). Most importantly, risk of UTI mortality increases with age, approaching 1/1000 population by age 80 (Yang et al., 2022).

Among older women and men, aged-associated factors, diabetes, hospitalization, placement of a urinary catheter, and a history of UTI, are associated with increased UTI risk (Foxman, 2014). Risk factors specific to men include urinary obstruction due to prostate enlargement associated with aging. Several studies have identified obesity as a UTI risk factor (Ribera et al., 2006; Semins et al., 2012; Seo et al., 2021), with one study showing the association to be independent of diabetes (Saliba et al., 2013). In all ages, sexual activity is associated with UTI risk (Rowe & Juthani-Mehta, 2013). However, the extent that these factors explain the increased UTI risk among older populations is unclear. Further, it is uncertain whether healthy habits, such as regular exercise, and urination habits reduce UTI risk among older individuals.

Constipation has been associated with UTI in children (Lorenzo et al., 2020a), and with urinary incontinence among women aged 18 and older (Baykuş & Yenal, 2020). The mechanism may be mechanical; anorectal and urodynamics showed that constipated young women have an increased bladder capacity and reduction in bladder and rectum sensitivity compared to that of women without constipation (Bannister et al., 1988). Further, a prospective Israeli study conducted among ambulatory patients aged 65 to 89 with chronic constipation and lower urinary tract symptoms found that treating constipation reduced urinary symptoms and residual urine volume (Charach et al., 2001). However, we found no studies reporting an association between constipation and UTI among adult women or men.

To describe the epidemiology and gain insight into associations between known and potential UTI risk factors among women and men aged 50 and older we added a questionnaire battery to the monthly Foresight 50+ Consumer Omnibus monthly survey during October, 2023. Results of this population-based study of 1034 Americans aged 50 years and older will be a resource for clinicians and stimulate future research on UTI etiology among older populations.

## Methods

### Study population

A general population sample of U.S. adults aged 50 and older was selected from NORC’s Foresight 50+ Panel, excluding any panelist who completed the previous wave. The study was fielded Oct 12 through Oct 16, 2023.

Funded and operated by NORC at the University of Chicago, Foresight 50+ is a probability-based panel designed to be representative of the U.S. household population age 50 or older. During the initial recruitment phase of the panel, randomly selected U.S. households were sampled with a known, non-zero probability of selection from the NORC National Sample Frame or a secondary national address frame, both with over 97% coverage of all U.S. addresses, and then contacted by U.S. mail, email, telephone, or field interviewers (face to face). Households were screened for having at least one adult age 50 and older. The panel provides sample coverage of approximately 97% of the U.S. household population. Those excluded from the sample include people with P.O. Box only addresses, some addresses not listed in the United States Postal Service Delivery Sequence File, and some newly constructed dwellings population. The panel also excludes people who live in some institutional types of settings, such as nursing facilities or nursing homes, depending on how addresses are listed for the facility. For panel sample selection, National Frame segments are stratified into six sampling strata based on the race/ethnicity and age composition of each segment. Areas with a higher concentration of young adults, Hispanics, and non-Hispanic African-Americans are oversampled to improve their representation in the panel.

The NORC study protocol was deemed to be exempt by the NORC Institutional Review Board (IRB00000967), under its Federalwide Assurance #FWA00000142 and the University of Michigan Medical School Institutional Review Board under its Federalwide Assurance #FWA00004969.

### Questionnaire

We added 24 questions specific to UTI to the NORC panel. These questions assessed UTI history, UTI diagnosis in the previous 12 months, frequency of an overnight hospital stay in the previous 12 months, current use of cranberry and estrogen products to prevent UTI, urination habits, sexual activity, and presence of urinary incontinence and constipation. As part of the NORC regular survey, we had access to sociodemographics, self-assessed overall health, exercise, height and weight. Most respondents completed the survey via a web interview (92%); the remainder completed the interview via phone. No question was skipped or missed by more than 4.2% of respondents, and for most questions the value was ≤ 1%.

### Analysis

All percents, Odds Ratios (OR) and Wald Confidence Limits (CL) were weighted using the probability of selection from the sampling frame to sample housing units for Foresight 50+ and adjusted to account for unknown eligibility and nonresponse to represent the U.S. population aged 50 and older defined by age, gender, race/ethnicity, and education.

The primary study outcome was self-reported UTI treated in the previous 12 months defined as a positive response to the question “Has a doctor ever told you that you had a urinary tract infection, cystitis or pyelonephritis?” and a response of 1 or more to the follow-up question “During the past 12 months, how many times have you had a urinary tract infection diagnosed and treated by a doctor?”. Results are presented as the 12-month incidence rate of UTI for each category of the exposure variables.

For descriptive analyses, numeric responses to exposure variables were categorized based on the overall distribution using quantiles. Categories for questions with responses assessed using predetermined categories were collapsed when samples sizes were small and the proportions with outcome in adjacent categories were similar. Differences in UTI incidence between categories were assessed using the Chi Square test. Associations were estimated using Rate Ratios (RR) and 95% confidence intervals.

After a thorough descriptive analysis, including assessment for confounding and interactions among variables, we fit multivariate models predicting UTI incidence to estimate ORs and Wald 95% CL for each selected variable adjusted for age. We fit a final model to better explain the novel association between constipation and UTI risk that included previously described UTI risk factors: age, gender, BMI>30 and an overnight hospital stay.

## Results

In this sample of 1074 Americans aged 50 and older participating in a probability-based panel designed to be representative of the U.S. household population age 50 or older, lifetime prevalence of self-reported healthcare provider diagnosed UTI was 49.7% among women and 12.7% among men. The self-reported incidence during the previous 12 months was 19.8% among women and 6.4% among men. Lifetime prevalence (Supplemental figure 1) and yearly incidence (Figure 1) varied by age, race/ethnic group, marital status and U.S. geographic region among women and men. Except for those aged 80 and older, the prevalence and incidence by each characteristic were higher among women than men.

**Figure 1:**
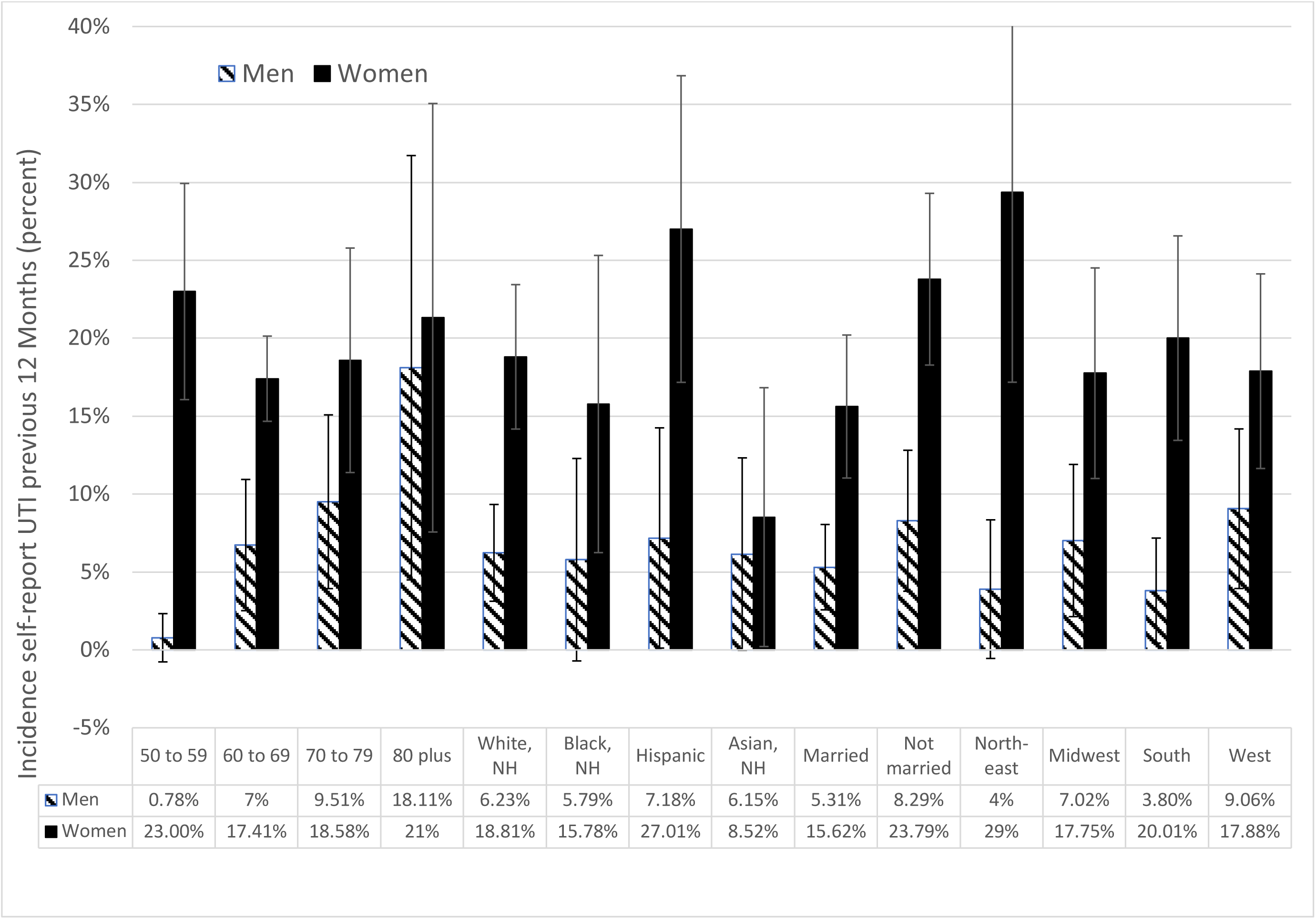
Self-reported incidence of healthcare provider diagnosed urinary tract infection, cystitis or pyelonephritis during the previous 12 months among 1074 Americans 50 and older in October 2023 by selected demographics. Final weighted data are representative of the U.S. household population age 50 or older. Error bars are +/-2 x the standard error of weighted percentage.

Incidence was highest among women aged 50 to 59 (23%) and lowest among women aged 60 to 69 (17%) after which it increased peaking again at age 80. Men’s incidence was lowest at ages 50 to 59 and increased thereafter. At ages 80 and older, incidence was 21% among women and 18% among men. Incidence varied significantly by racial/ethnic group among women (p=0.02), with highest incidence among Hispanic women and lowest among Non-Hispanic Asian women. Incidence was not statistically significantly different by racial/ethnic groups among men. Men and women who were married had lower incidence than those who were widowed, divorced, separated, never married or living with a partner. By region, incidence was highest in the Northeast for women (29%) and in the West for men (9%). Among those with a UTI in the previous 12 months, 10% of women and 7% of men reported 3 or more UTIs in the previous year. The distribution of number of self-reported UTI in the previous 12 months fits an exponential distribution (Figure 2).

**Figure 2:**
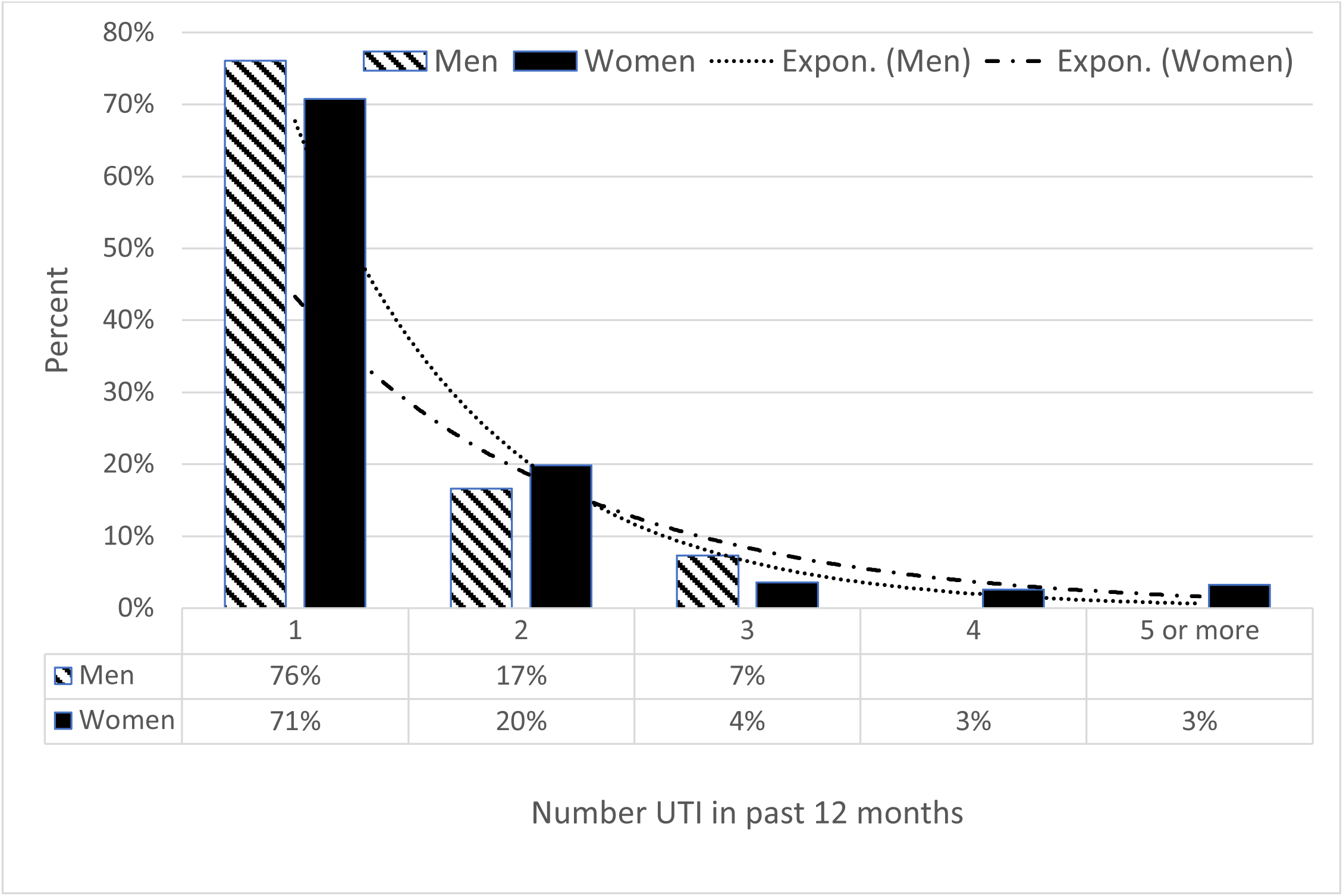
Number of self-reported healthcare provider diagnosed urinary tract infection, cystitis or pyelonephritis among 1074 Americans 50 and older in the previous 12 months in October 2023 by gender. Final weighted data are representative of the U.S. household population age 50 or older.

After adjustment for age several variables were significantly associated with reporting a UTI in the previous 12 months among women and men. UTI incidence was lowest among women and men reporting excellent or very good overall health. Women and men who had a body mass index (BMI) of 30 or more, an overnight hospital stay, did not exercise, and used tobacco every day were significantly more likely to report a UTI in the previous 12 months (Table 1). Leaking urine, delaying urination, and urinating 3 or more times per night were also associated with UTI incidence among women and men. There was a dose response relationship between increasing frequency of constipation and increasing UTI incidence among women and men.

**Table 1:**
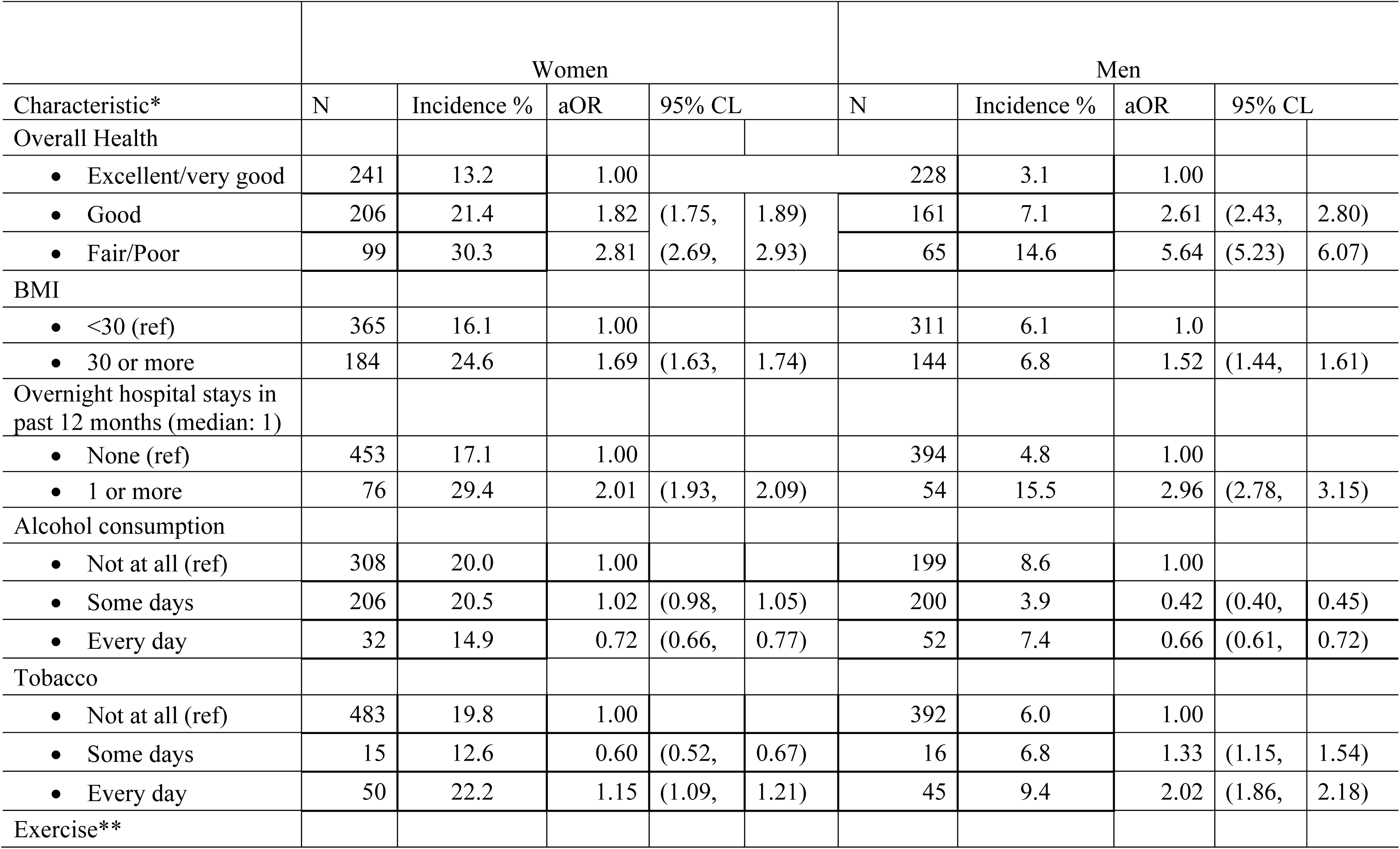
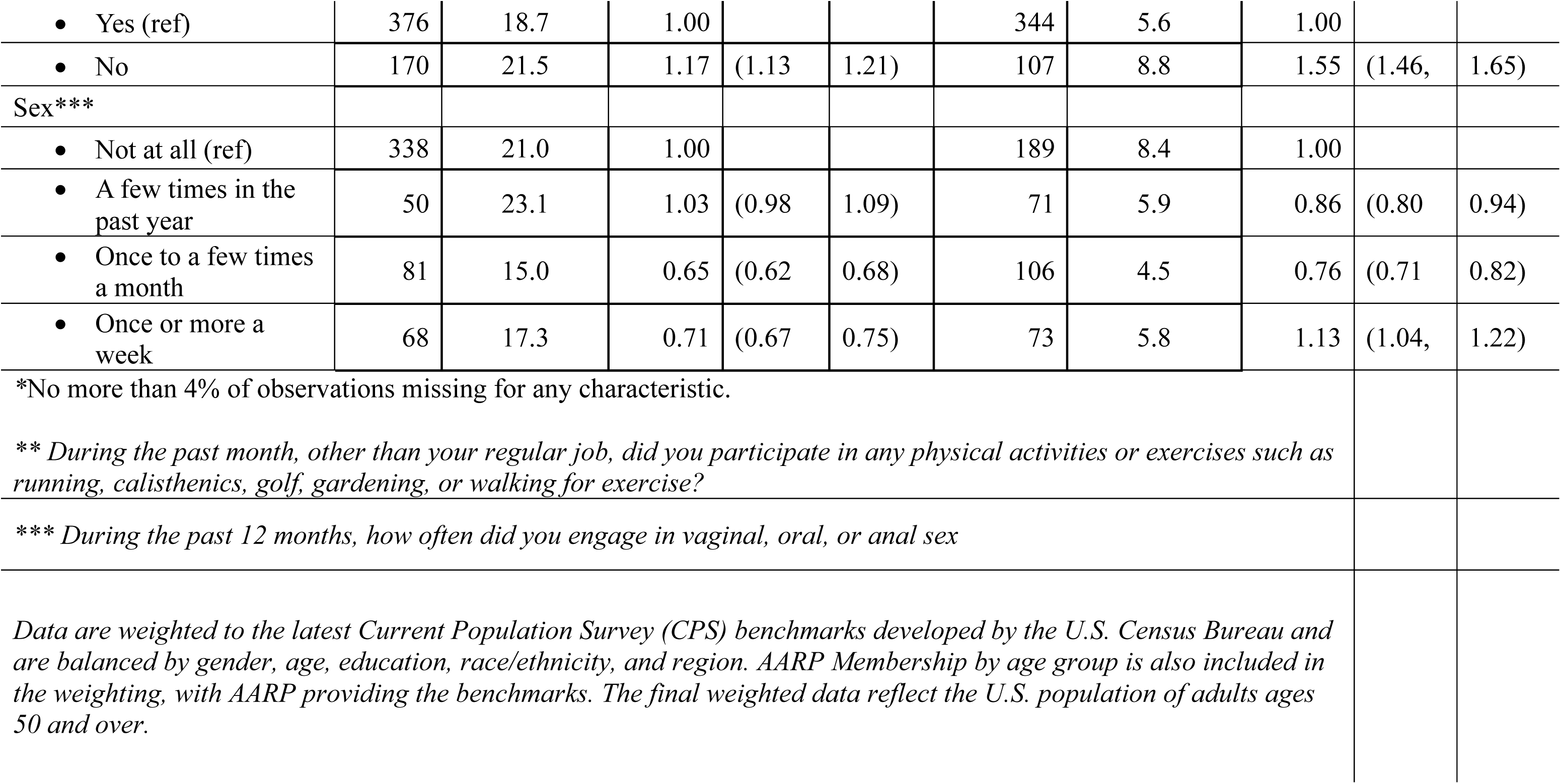
Urinary tract infection annual incidence rates (weighted to reflect the US population) and age-adjusted associations with selected measures of health by gender among 1074 Americans aged 50 and older. Percent with UTI in each category, Odds ratios (ORs) and Wald Confidence Limits (CL) weighted to represent the U.S. population. Results from a probability-based panel designed representative of the U.S. household population age 50 or older, October, 2023.

Overall, 41% of women and 23% of men reported they experienced constipation sometimes, often or almost always. There were no statistically significant differences in constipation by race/ethnic group, marital status or BMI, but self-report of some, often or always constipation significantly increased among those who delayed urination or who had an on overnight stay (Figure 3). UTI incidence increased with constipation frequency among women and men (Table 2). After adjusting for age, gender, BMI, and an overnight hospital stay those reporting often or always experience constipation were 5.48 times (95% CL: 5.13,5.84) more likely than those reporting never having constipation to report a UTI in the previous 12 months (Table 3).

**Figure 3:**
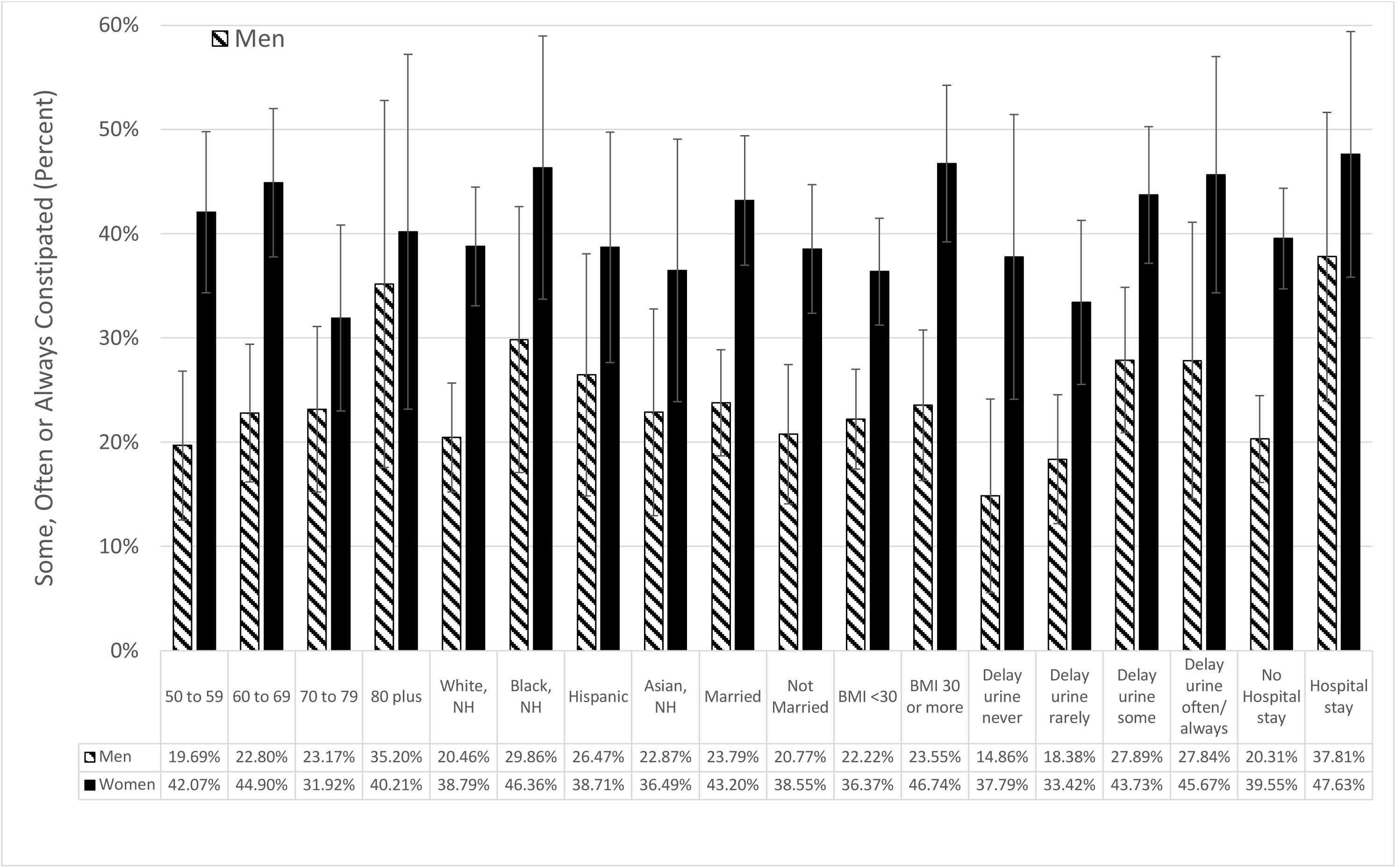
Self-reported prevalence of sometimes, often or always experiencing constipation among 1074 Americans 50 and older by selected demographics. Percents weighted to represent the U.S. population. Results from a probability-based panel designed to be representative of the U.S. household population age 50 or older. October, 2023. Error bars are +/-2 x the standard error of weighted percentage.

**Table 2:**
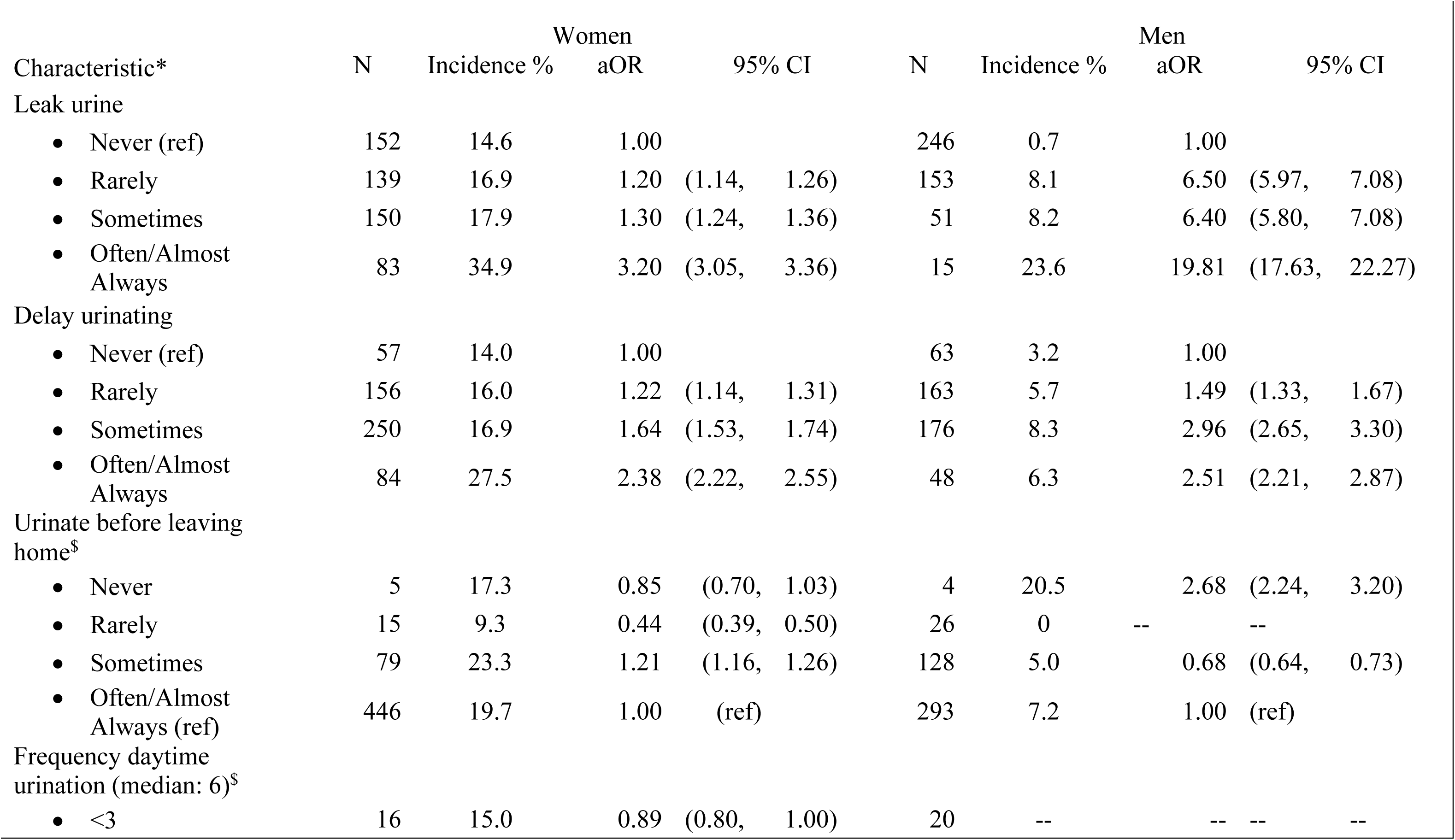
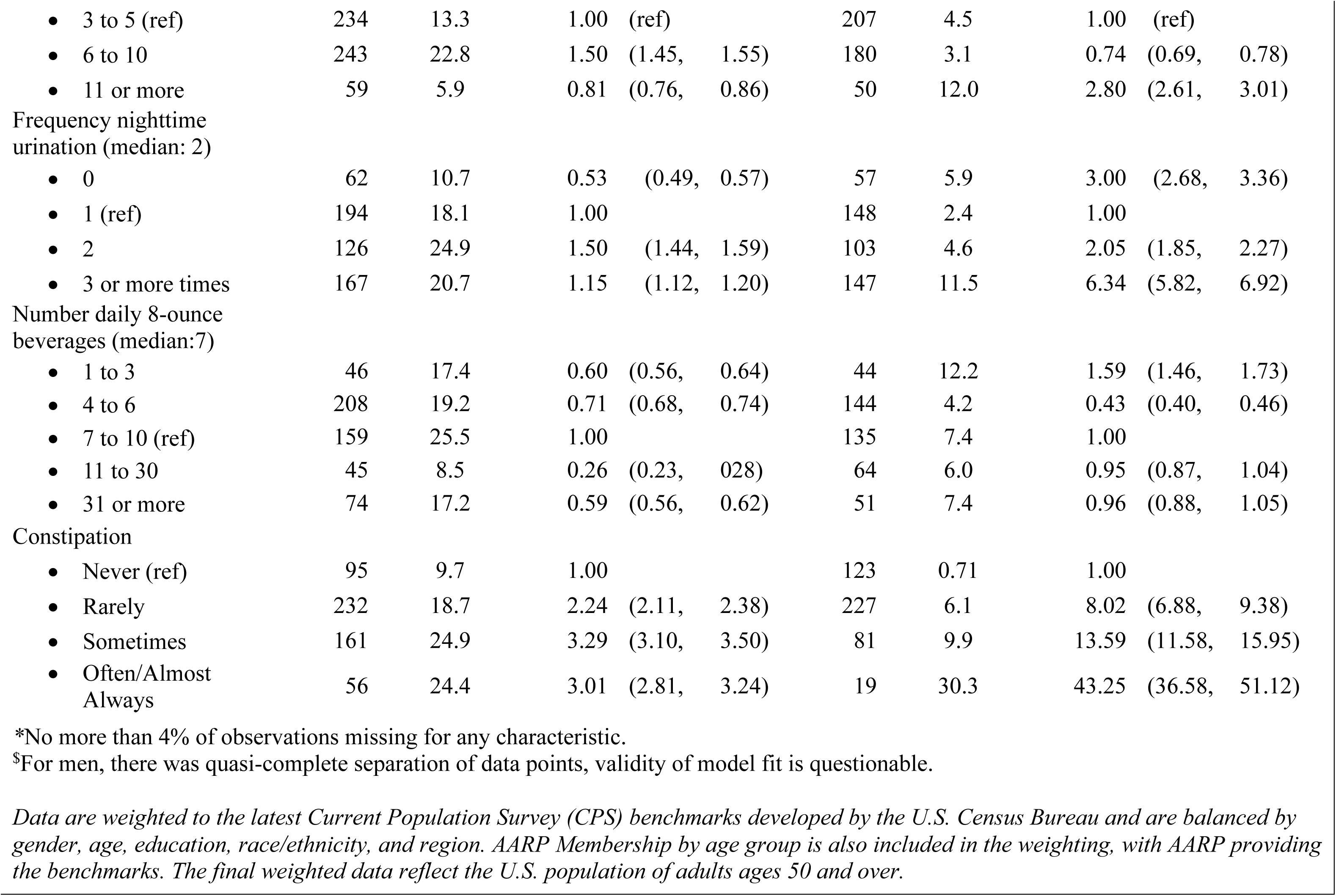
Urinary tract infection annual incidence rates (weighted to reflect the US population) and age-adjusted associations with selected measures of health behaviors by gender among 1074 Americans aged 50 and older. Percent with UTI in each category, Odds ratios (ORs) and Confidence Limits (CL) weighted to represent the U.S. population. Results from a probability-based panel representative of the U.S. household population age 50 or older, October, 2023.

**Table 3:** Adjusted Odds ratios (aORs) and Wald 95 percent confidence limits (95% CL) from a logistic model predicting urinary tract infection incidence by gender, age, biomass index (BMI) and self-reported constipation. Odds ratios (ORs) and Confidence Limits (CL) weighted to represent the U.S. population. Results from a probability-based panel representative of the U.S. household population age 50 or older, October, 2023.

|  |  | Model including Women and Men |  |  |  |
| --- | --- | --- | --- | --- | --- |
| Variable |  | Beta | p value | aOR | 95%CI |
| Gender | Male |  |  | 1.00 | (ref) |
|  | Female | 1.1315 | <0.0001 | 3.10 | (3.00, 3.21) |
| Age | 50-59 | 0.0145 | 0.438 | 1.02 | (0.98, 1.05) |
|  | 60-69 | -- |  | 1.00 | (ref) |
|  | 70-79 | 0.3315 | <0.0001 | 1.39 | (1.34, 1.45) |
|  | 80+ | .6958 | <0.0001 | 2.01 | (1.90, 2.12) |
| BMI | <30 |  |  | 1.00 | (ref) |
|  | 30 or more | 0.2850 | <0.0001 | 1.33 | (1.29, 1.37) |
| Overnight hospital stay | no |  |  | 1.00 | (ref) |
|  | yes | 0.7104 | <0.0001 | 2.04 | (1.96, 2.11) |
| Constipation | never |  |  | 1 | (ref) |
|  | rare | 0.9484 | <0.0001 | 2.58 | (2.44, 2.73) |
|  | sometimes | 1.3064 | <0.0001 | 3.69 | (3.49, 3.91) |
|  | often/always | 1.7003 | <0.0001 | 5.48 | (5.13, 5.84) |

*Data are weighted to the latest Current Population Survey (CPS) benchmarks developed by the U.S. Census Bureau and are balanced by gender, age, education, race/ethnicity, and region. AARP Membership by age group is also included in the weighting, with AARP providing the benchmarks. The final weighted data reflect the U.S. population of adults ages 50 and over.*

In this population, 32% reported experiencing constipation sometimes, often or always. We used this percent, and the adjusted OR for experiencing constipation sometimes compared to never (3.69) to estimate the population attributable fraction [(proportion with constipation * (OR-1)(/proportion with constipation * (OR-1) +1))]. We used the OR for sometimes versus never rather than the OR for frequently or always (5.48) to be conservative. Using those values, we estimated that the proportion of UTI among those 50 and older that might be attributable to constipation as 46% [(.32*(3.69-1))/((.32*(3.69-1)+1)].

## Discussion

Constipation has previously been identified as a UTI risk factor among children (Lorenzo et al., 2020b). Here we present the first report of an association between constipation and UTI among older women and men; notably the strength of this association increased as constipation increased in frequency and the association remained after adjustment for other known UTI risk factors: age, gender, BMI >30, and an overnight hospital stay. Constipation is common in older populations: 41% of women and 23% of men in our survey reported sometimes, often or always experiencing constipation.

There is reason to believe that treating constipation might reduce UTI incidence: a prospective study among ambulatory older men and women showed that treating constipation reduced urinary symptoms and residual urine volume (Charach et al., 2001). Thus, treating constipation might also reduce UTI occurrence. Based on our results, we estimated that 46% of UTI among those 50 and older might be attributable to constipation. Using our estimates of UTI incidence and the 2020 U.S. population of 65 and older (55,792,501) there are almost 6 million (10.6% x 55,792,501=5,914,005) UTIs diagnosed and treated by a healthcare provider among Americans eligible for Medicare. Therefore, treating constipation might have results in a significant reduction in societal and personal costs.

Reducing the number UTI would also reduce antibiotic prescriptions. Antibiotic resistance arises and spreads in response to antibiotic treatment. The selection pressure that occurs from treating 73 people for a 5-day course with the same antibiotic is roughly equivalent in terms of probability of resistance emerging as treating 1 person for 365 days. Between 80 and 90% of uncomplicated UTIs are caused by *Escherichia coli*. In 2022, the Global Antimicrobial Resistance and Use Surveillance System reported that resistance to sulfonamide and trimethoprim, fluoroquinolones, and some third-generation cephalosporins among *E.coli* UTIs exceeded 20% (World Health Organization, 2022). Among *E. coli* UTI causing uncomplicated UTI treated at Kaiser Permanente Southern California between January 2016 and December 2021 resistance to 1, 2, 3, and 4 antibiotic classes was found in 19%, 18%, 8%, and 4% of isolates, respectively; 1% were resistant to ≥5 antibiotic classes, and 50% were resistant to none (Ku et al., 2023).

Our estimates of UTI incidence are somewhat higher than found in the analysis of NHANES-III (1988 to 1994) data (Griebling, 2005b, 2005a). At ages 80 and older, 21% of women and 18% men in our study reported a UTI in the previous year compared to 11.7% women and 7.7% of men aged 85 and older in NHANES-III. By contrast, our estimates of lifetime estimates were somewhat lower (women: 47.1% vs. 53.1%; men: 9.3% vs. 13.6%). The NHANES-III lifetime prevalences were estimated using data from adults18 and older; our data were limited to adults 50 and older. Recall of acute infections may be poor: in our study women aged 50 to 59 reported more UTIs in the previous 12 months then women aged 60 to 69. Previous clinical estimates of risk of recurring UTI among healthy women are 2 to 5% (Nicolle, 2011), somewhat lower than the 10% among women and 7% men reported here for those 50 and older. We found no published estimates specific to older populations or for men.

Engaging in sexual activity is associated with UTI risk in younger populations (Brown & Foxman, 2000); at least one large study among post-menopausal women suggested this may also be the case in older women (Moore et al., 2008). We found no studies supporting or refuting an association of sexual activity and UTI among men. In our study most men and women reported no sexual of activity during the previous year; after adjusting for age, there was no association between more frequent sexual intercourse and UTI. We note, however, that UTI in the past year was higher among women and men who were not married, and who possibly engaged in intimate contact (including sexual intercourse) with one or more partners. This is consistent with other studies supporting transmission of uropathogens with intimate contact (Foxman et al., 2002).

We found only one report regarding cigarette smoking and UTI risk. This large case control study (601,352 UTI cases, 1,303,455 controls) identified participants from electronic health records; there was no association with cigarette smoking when calculated using the raw data provided (Casey et al., 2021). Models were adjusted for cigarette smoking but the estimates for smoking were not included. However, cigarette smoking was associated with overactive bladder symptoms in a cross-sectional questionnaire study of Japanese study of 4756 women (Kawahara et al., 2020) and in a British study cigarette smoking was associated with secondary nocturnal enuresis (Madhu et al., 2017). It is possible that some of the self-reported UTIs in our study are other bladder conditions, or that the association we observed is explained by unmeasured confounders. Nonetheless, this association is worth further investigation.

After adjusting for age, gender, an overnight stay in hospital, and constipation we identified a significant association between BMI ≥ 30 and UTI. No previous studies accounted for constipation. However, there are previous reports of associations of UTI with BMI. An Israeli record-review study of 110,736 women and 42,703 men aged 18 and older found an increase in UTI incidence with increasing BMI with the greatest effect sizes for men at highest levels of BMI (Hazard ratio for BMI > 50 kg/m^2^ compared to <25 kg/m^2^ : Men: 2.38; (95%CI: 1.40-4.03); Women: 1.39 (95% CI:1.03 -1.52), adjusted for age, history of diabetes, serum 25(OH)D quartile (Saliba et al., 2013). In a Korean community-based cohort study (n=4,926; median age 59; range 48 to 80) there was the hazard ratio (HR) for those with BMI 30 or over compared to normal weight after adjusting for gender, age, residence, living with spouse, current smoking, current drinking and diabetes was 1.66 (1.06, 2.61) (Seo et al., 2021). The adjusted HR was greater for men (4.37; 95% CI: 1.11, 17.15) than for women (1.58; 95% CI 0.98-2.55).

Self-reported ratings of overall health have been associated in multiple studies with several chronic diseases (H. Yang et al., 2021) and overall mortality (Lorem et al., 2020). Thus, our report that women and men who self-assessed their overall health as fair or poor were more likely to report UTI in the previous 12 months than those rating their health as very good/excellent is consistent with previous reports. It is also consistent with the significant association with an overnight hospital stay in the previous 12 months, which is a known UTI risk factor. As self-rated health is derived from how the individual assesses their health problems, general physical functioning and the ‘healthiness’ of their own behaviors (Quesnel-Vallée, 2007) this first report of an association of self-rated overall health with UTI may stimulate further UTI research among the elderly.

Our study has several strengths. The sample size is large, and the results are weighted to represent the U.S. household population aged 50 and older. Further, it one of very few studies to identify risk factors among women and men. However, for variables such as urination habits, the results should be interpreted with caution. We did not have the date of the UTI, which is critical for evaluating whether behaviors that might change in response to UTI such as delaying or more frequently urinating, or increased beverage consumption might have a causal relationship.

Our measure of constipation was based on a single question “How often are you constipated? Examples include stools that are separate hard lumps like nuts that are difficult to pass or sausage-shaped but lumpy and difficult to pass” which only captures part of the Rome criteria for functional constipation (Barberio et al., 2021). A final limitation is that UTI was assessed by self-report of diagnosis and treatment by a healthcare provider. It is possible that respondents may not have accurately remembered their diagnosis. Further, UTI is usually diagnosed among outpatients based on symptoms or symptoms and urinalysis without benefit of culture. Even if there is a positive urinalysis or culture, asymptomatic bacteriuria is common – especially among older populations, and the symptoms are not specific to UTI, so some misclassification is likely. Despite these limitations, the associations between UTI and constipation are strong, occur in women and men in a dose response manner and are biologically plausible: constipation negatively affects bladder and rectum sensitivity and increases residual volume.

## Data Availability

All data produced in the present work are contained in the manuscript

## Acknowledgements

The authors thank Anna Cronenwett for administrative assistance. This project was funded by the Hunein F. and Hilda Maassab Endowment (BF) and the Haas Family.

## Acknowledgements

This project was generously supported by funds from the Hass family and the Maassab Endowment fund.

---

**Supplemental Figure 1:**
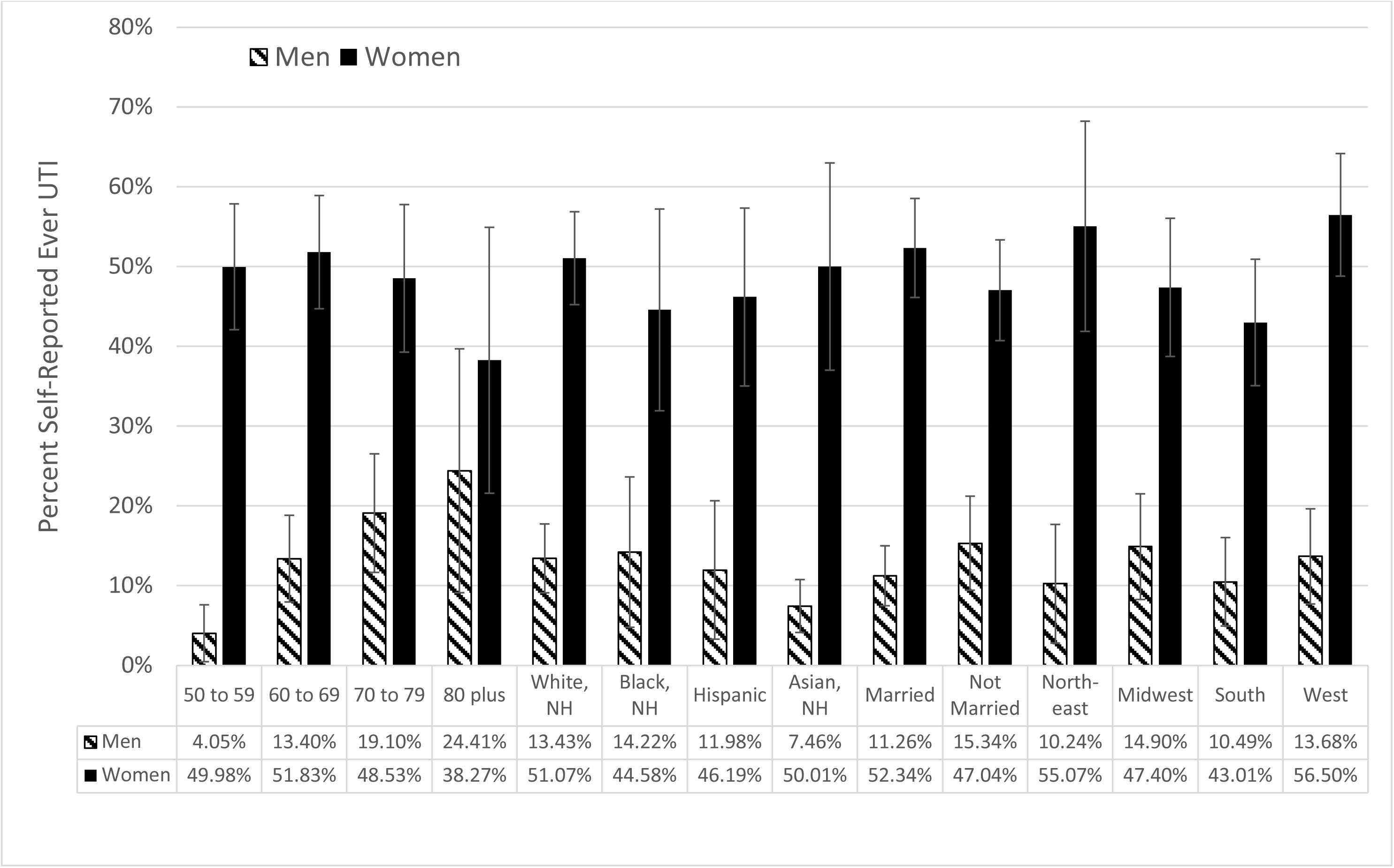
Self-reported lifetime prevalence of healthcare provider diagnosed urinary tract infection, cystitis or pyelonephritis among 1074 Americans 50 and older by selected demographics. Results from a probability-based panel designed to be representative of the U.S. household population age 50 or older. October, 2023. Error bars are +/-2 x the standard error of weighted percentage.

## References

1. Arinzon, Z., Shabat, S., Peisakh, A., & Berner, Y. (2012). Clinical presentation of urinary tract infection (UTI) differs with aging in women. Archives of Gerontology and Geriatrics, 55(1), 145–147. 10.1016/J.ARCHGER.2011.07.012

2. Bannister, J. J., Lawrence, W. T., Smith, A., Thomas, D. G., & Read, N. W. (1988). Urological abnormalities in young women with severe constipation. Gut, 29(1), 17–20. 10.1136/GUT.29.1.17

3. Barberio, B., Judge, C., Savarino, E. V., & Ford, A. C. (2021). Global prevalence of functional constipation according to the Rome criteria: a systematic review and meta-analysis. *The Lancet*. Gastroenterology & Hepatology, 6(8), 638–648. 10.1016/S2468-1253(21)00111-4

4. Baykuş, N., & Yenal, K. (2020). Prevalence of urinary incontinence in women aged 18 and over and affecting factors. Journal of Women & Aging, 32(5), 578–590. 10.1080/08952841.2019.1682923

5. Brown, P. D., & Foxman, B. (2000). Pathogenesis of Urinary Tract Infection: the Role of Sexual Behavior and Sexual Transmission. Current Infectious Disease Reports, 2(6), 513–517. 10.1007/S11908-000-0054-4

6. Bruxvoort, K. J., Bider-Canfield, Z., Casey, J. A., Qian, L., Pressman, A., Liang, A. S., Robinson, S., Jacobsen, S. J., & Tartof, S. Y. (2020). Outpatient Urinary Tract Infections in an Era of Virtual Healthcare: Trends From 2008 to 2017. Clinical Infectious Diseases : An Official Publication of the Infectious Diseases Society of America, 71(1), 100–108. 10.1093/CID/CIZ764

7. Cairns C, Kang K. National Hospital Ambulatory Medical Care Survey: 2020 emergency department summary tables. DOI: 10.15620/cdc:121911.]

8. Casey, J. A., Rudolph, K. E., Robinson, S. C., Bruxvoort, K., Raphael, E., Hong, V., Pressman, A., Morello-Frosch, R., Wei, R. X., & Tartof, S. Y. (2021). Sociodemographic Inequalities in Urinary Tract Infection in 2 Large California Health Systems. Open Forum Infectious Diseases, 8(6). 10.1093/OFID/OFAB276

9. Charach, G., Greenstein, A., Rabinovich, P., Groskopf, I., & Weintraub, M. (2001). Alleviating constipation in the elderly improves lower urinary tract symptoms. Gerontology, 47(2), 72–76. 10.1159/000052776

10. Foxman, B. (2014). Urinary tract infection syndromes: occurrence, recurrence, bacteriology, risk factors, and disease burden. Infectious Disease Clinics of North America, 28(1), 1–13. 10.1016/J.IDC.2013.09.003

11. Foxman, B., Manning, S. D., Tallman, P., Bauer, R., Zhang, L., Koopman, J. S., Gillespie, B., Sobel, J. D., & Marrs, C. F. (2002). Uropathogenic Escherichia coli are more likely than commensal E. coli to be shared between heterosexual sex partners. American Journal of Epidemiology, 156(12), 1133–1140. 10.1093/AJE/KWF159

12. Gharbi M, Drysdale JH, Lishman H, Goudie R, Molokhia M, Johnson AP, Holmes AH, Aylin P (2019). Antibiotic management of urinary tract infection in elderly patients in primary care and its association with bloodstream infections and all cause mortality: population-based cohort study. British Medical Journal, 364:l525. doi: 10.1136/bmj.l525.

13. Griebling, T. L. (2005a). Urologic diseases in america project: trends in resource use for urinary tract infections in men. The Journal of Urology, 173(4), 1288–1294. 10.1097/01.JU.0000155595.98120.8E

14. Griebling, T. L. (2005b). Urologic diseases in America project: trends in resource use for urinary tract infections in women. The Journal of Urology, 173(4), 1281–1287. 10.1097/01.JU.0000155596.98780.82

15. Kawahara, T., Ito, H., Yao, M., & Uemura, H. (2020). Impact of smoking habit on overactive bladder symptoms and incontinence in women. International Journal of Urology : Official Journal of the Japanese Urological Association, 27(12), 1078–1086. 10.1111/IJU.14357

16. Ku, J. H., Bruxvoort, K. J., Salas, S. B., Varley, C. D., Casey, J. A., Raphael, E., Robinson, S. C., Nachman, K. E., Lewin, B. J., Contreras, R., Wei, R. X., Pomichowski, M. E., Takhar, H. S., & Tartof, S. Y. (2023). Multidrug Resistance of Escherichia coli From Outpatient Uncomplicated Urinary Tract Infections in a Large United States Integrated Healthcare Organization. Open Forum Infectious Diseases, 10(7). 10.1093/OFID/OFAD287

17. Lorem, G., Cook, S., Leon, D. A., Emaus, N., & Schirmer, H. (2020). Self-reported health as a predictor of mortality: A cohort study of its relation to other health measurements and observation time. Scientific Reports, 10(1). 10.1038/S41598-020-61603-0

18. Lorenzo, A. J., Rickard, M., & Santos, J. Dos. (2020a). The role of bladder function in the pathogenesis and treatment of urinary tract infections in toilet-trained children. *Pediatric Nephrology (Berlin*, Germany), 35(8), 1395–1408. 10.1007/S00467-019-4193-6

19. Lorenzo, A. J., Rickard, M., & Santos, J. Dos. (2020b). The role of bladder function in the pathogenesis and treatment of urinary tract infections in toilet-trained children. *Pediatric Nephrology (Berlin*, Germany), 35(8), 1395–1408. 10.1007/S00467-019-4193-6

20. Madhu, C. K., Hashim, H., Enki, D., & Drake, M. J. (2017). Risk factors and functional abnormalities associated with adult onset secondary nocturnal enuresis in women. Neurourology and Urodynamics, 36(1), 188–191. 10.1002/NAU.22912

21. Moore, E. E., Hawes, S. E., Scholes, D., Boyko, E. J., Hughes, J. P., & Fihn, S. D. (2008). Sexual intercourse and risk of symptomatic urinary tract infection in post-menopausal women. Journal of General Internal Medicine, 23(5), 595–599. 10.1007/s11606-008-0535-y

22. Nicolle, L. E. (2011). Update in adult urinary tract infection. Current Infectious Disease Reports, 13(6), 552–560. 10.1007/S11908-011-0212-X

23. Quesnel-Vallée, A. (2007). Self-rated health: caught in the crossfire of the quest for “true” health? International Journal of Epidemiology, 36(6), 1161–1164. 10.1093/IJE/DYM236

24. Ribera, M. C., Pascual, R., Orozco, D., Pérez Barba, C., Pedrera, V., & Gil, V. (2006). Incidence and risk factors associated with urinary tract infection in diabetic patients with and without asymptomatic bacteriuria. European Journal of Clinical Microbiology & Infectious Diseases : Official Publication of the European Society of Clinical Microbiology, 25(6), 389–393. 10.1007/S10096-006-0148-5

25. Rowe, T. A., & Juthani-Mehta, M. (2013). Urinary tract infection in older adults. Aging Health, 9(5), 519–528. 10.2217/AHE.13.38

26. Saliba, W., Barnett-Griness, O., & Rennert, G. (2013). The association between obesity and urinary tract infection. European Journal of Internal Medicine, 24(2), 127–131. 10.1016/J.EJIM.2012.11.006

27. Santo L, Kang K. National Ambulatory Medical Care Survey: 2019 National Summary Tables. Available from: DOI: 10.15620/cdc:123251.

28. Semins, M. J., Shore, A. D., Makary, M. A., Weiner, J., & Matlaga, B. R. (2012). The impact of obesity on urinary tract infection risk. Urology, 79(2), 266–269. 10.1016/J.UROLOGY.2011.09.040

29. Seo, S. H., Jeong, I. S., & Lee, E. J. (2021). Impact of Obesity on Urinary Tract Infections in Korean Adults: Secondary Data Analysis Using Community-Based Cohort Study. Journal of Korean Academy of Nursing, 51(2), 150–161. 10.4040/JKAN.20228

30. Strassle, P. D., Sickbert-Bennett, E. E., Klompas, M., Lund, J. L., Stewart, P. W., Marx, A. H., Dibiase, L. M., & Weber, D. J. (2019). Incidence and risk factors of non-device-associated urinary tract infections in an acute-care hospital. Infection Control and Hospital Epidemiology, 40(11), 1242–1247. 10.1017/ICE.2019.241

31. Sulham, K., & Hammelman, E. (2021). 1416. Medicare Spending on Urinary Tract Infections: A Retrospective Database Analysis. Open Forum Infectious Diseases, 8(Suppl 1), S793. 10.1093/OFID/OFAB466.1608

32. World Health Organization. (2022). Global Antimicrobial Resistance and Use Surveillance System (GLASS) Report 2022. In World Health Organization (Issue 8.5.2017). https://www.who.int/publications/i/item/9789240062702

33. Yang, H., Deng, Q., Geng, Q., Tang, Y., Ma, J., Ye, W., Gan, Q., Rehemayi, R., Gao, X., & Zhu, C. (2021). Association of self-rated health with chronic disease, mental health symptom and social relationship in older people. Scientific Reports, 11(1). 10.1038/S41598-021-94318-X

34. Yang, X., Chen, H., Zheng, Y., Qu, S., Wang, H., & Yi, F. (2022). Disease burden and long-term trends of urinary tract infections: A worldwide report. Frontiers in Public Health, 10. 10.3389/FPUBH.2022.888205

